# BA.2 evasion of vaccine induced binding and functional non-neutralizing antibodies

**DOI:** 10.1101/2022.02.25.22271511

**Authors:** Yannic C Bartsch, Deniz Cizmeci, Jaewon Kang, Hailong Gao, Wei Shi, Abishek Chandrashekar, Ai-ris Y Collier, Bing Chen, Dan H Barouch, Galit Alter

**Affiliations:** Ragon Institute of MGH, MIT, and Harvard; Cambridge, MA, USA; Division of Molecular Medicine, Boston Children’s Hospital, Harvard Medical School, Boston, MA, USA; Beth Israel Deaconess Medical Center, Boston, MA, USA

## Abstract

The number of mutations in the omicron (B1.1.529) BA.1 variant of concern (VOC) led to an unprecedented evasion of vaccine induced immunity. However, despite the remarkable rise in global infections, severe disease and death did not increase proportionally, linked to persistent recognition of BA.1 by T cells and non-neutralizing opsonophagocytic antibodies. Yet, the emergence of a new sublineage, BA.2, that is more transmissible compared to BA.1, despite relatively preserved neutralizing antibody responses, has raised the possibility that BA.2 may evade other vaccine induced responses that may be key to protection against infection and disease. Here we comprehensively profiled the BNT162b2 vaccine induced response to several VOCs including the omicron BA.1 and BA.2, after the primary vaccine series, 8 months following vaccination, and after a boost. While vaccine induced immune responses were compromised against both Omicron sublineages, vaccine induced antibody isotype titers, FcγR3a- and FcγR3b-receptor binding levels, and non-neutralizing opsonophagocytic functions were significantly attenuated to the omicron BA.2 Spike compared to the BA.1 lineage. Conversely, FcγR2a and FcγR2b binding was elevated to BA.2 potentially contributing to persistent protection against severity of disease. These data point to an attenuation of particular non-neutralizing antibody properties that may be key to protection against transmission, but the maintenance of others that may continue to confer protection against disease.

## Introduction

The rapid and perpetual evolution of SARS-CoV-2 continues to raise concerns related to the potential evolution of more pathogenic variants able to evade natural or vaccine induced immune responses. With each wave of variants of concern (VOCs), SARS-CoV-2 has acquired mutations that have increased infectivity, either via stabilization of the spike, enhanced binding to the angiotensin converting enzyme-2 (ACE-2), or via alterations in the viral fusion machinery (*1-3*). Evasion from neutralizing antibodies has progressed in parallel to this evolution, due to the natural accumulation of mutations near or around sites involved in infection and fusion (*4, 5*). The most recent variant of concern, omicron, has exhibited the greatest evolutionary leap, acquiring 36 mutations, deletions, or insertions in the Spike antigen, associated with significant evasion of vaccine induced neutralizing antibodies and increased infectivity. However, despite this remarkable increase in transmissibility, the omicron BA.1 variant has exhibited overall lower pathogenicity and disease (*5-8*).

Yet, a new omicron sublineage has recently emerged, the BA.2 lineage. While BA.2 Spike lacks 16 alterations characteristic to BA1 it has acquired 11 unique additional amino acid changes that are speculated to have increased BA.2 infectivity by 30% over BA.1 (*9*). The BA.2 lineage has now taken over the epidemic across Southeast Asia, Africa, Denmark, and is on the rise across Europe and the Americas (*10*). Like the parental BA.1, BA.2 evades natural or waning vaccine induced neutralizing antibodies equally but can be neutralized with boosting (*11, 12*).

While neutralizing antibodies show minimally changed reactivity to BA.2, we speculated that other immune mechanisms may lose potency against BA.2, enabling this lineage to spread more efficiently. In addition to neutralization, antibodies contribute to prevention of infection via their ability to leverage the innate immune response, including the recruitment of opsonophagocytic, via the recruitment of innate cellular function via Fc-receptors. Given the accumulating data pointing to a role for Fc-effector function in protection against severe COVID-19, monoclonal therapeutic efficacy, as well as vaccine mediated protection (*13-16*), here we aimed to define whether BA.2 evades these additional functions of the BNT16b2 vaccine induced humoral immune response at peak immunogenicity after the primary series, after 8 months, as well as after a boost. We observed significant differences in vaccine induced immunity to BA.1 and BA.2, marked by significantly lower antibody binding titers, selective loss of BA.2 FcγR3a and FcγR3b binding antibodies, as well as reduced neutrophil opsonophagocytic function to BA.2. Thus, despite the near identical neutralizing antibody responses to BA.2, vaccine induced Fc-effector functions are selectively compromised to BA.2, potentially marking a weakened mucosal opsonophagocytic response to this virus that may be key to attenuating mucosal transmission.

## Results

### Diminished isotype binding titers to BA.2

Despite nearly identical vaccine induced neutralizing antibody responses to the BA.1 and BA.2, the BA.2 sublineage exhibits a 30% increase in infectiousness and a potential increase in disease severity (*9*), calling into question whether this mutant may evade vaccine induced immunity in a manner that is distinct from the original omicron BA.1 variant. Thus, to begin we probed the ability of Pfizer BNT162b2 vaccine induced antibodies to bind across variants of concern (VOCs) including the wildtype/wuhan, Alpha, Beta, Delta, and Omicron BA.1 and BA.2 variants at peak immunogenicity (2 weeks after the primary first and second vaccine), after 8 months, and 2 weeks following a BNT162b2 boost (**Figure 1**). Binding IgM, IgA, and IgG responses were probed. Robust IgM binding titers were observed across most VOCs after the primary series, albeit response to BA.1 were lower and lowest to BA.2. IgM responses were rapidly lost over time and were not boosted against the beta, delta, or omicron Spikes (**Figure 1A**). A similar profile was observed in the IgA response (**Figure 1B**), marked by robust IgA levels to all VOCs to the D614G wildtype, alpha, beta, and delta variant, but lower responses to the omicron BA.1 and even lower responses to the BA.2. Moreover, the responses waned over the first 8 months proportionally to all variants, but most significantly for the omicron BA.2. However, all IgA responses boosted, albeit the omicron BA.1 and BA.2 responses never reached levels observed to the other VOCs. Moreover, BA.2 IgA responses remained significantly lower to those observed to BA.1. Finally, IgG responses showed a similar profile to IgA, with lower IgG binding tiers to the omicron BA.1 and BA.2, with the lowest responses to the BA.2 Spike (**Figure 1C**), IgG levels waned markedly for all VOCs, but most considerably to omicron binding IgG levels. All VOC IgG levels were boosted after the third dose of BNT162b2, although IgG to BA.2 did not boost proportionately.

**Figure 1:**
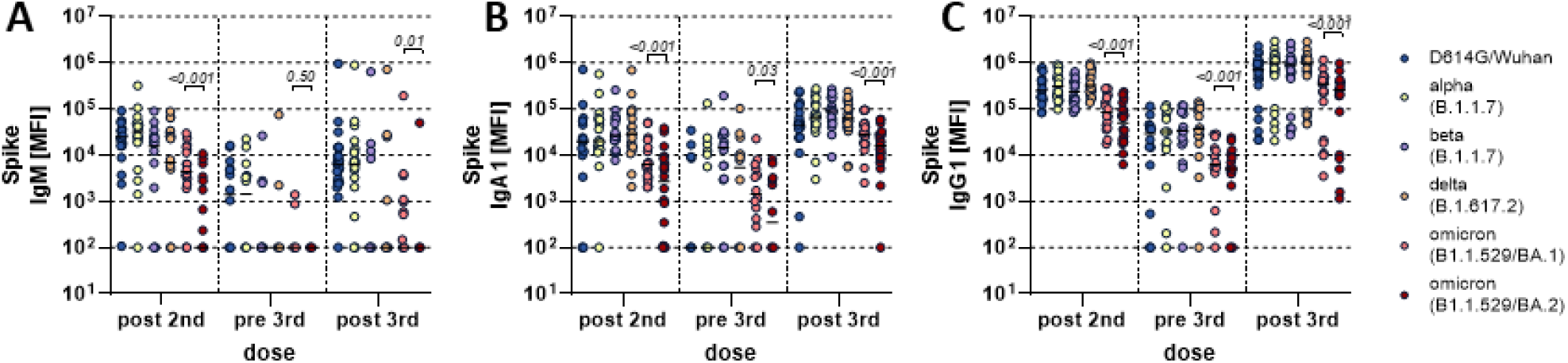
BNT162b2 induced antibody binding titer to different SARS-CoV-2 Spike variants of concern. Individuals received a three-dose regimen of the BNT162b2 vaccine. Samples were taken approx. 4 weeks after the second dose (post 2nd, n=18), before the third dose approx. 8 months after the second dose (pre 3rd, n=14) and approx. 4 weeks after the third dose (post 3rd, n=22). IgM (A), IgA1 (B) and IgG1 (C) binding titers to D614G (WT), Alpha (B.1.1.7), Beta (B.1.351), Delta (B.1.617.2) variants of concern, and Omicron (B.1.1.529) BA.1 and BA.2 subvariants Spike were measured by Luminex. The average value of technical replicates is shown. The data was corrected for background and negative values were set to 100 for graphing purposes. A two-sided paired Wilcoxon test with a Benjamini-Hochberg post-test correcting for multiple comparisons was used to test for statistical differences between BA1 and BA2 titers, respectively. P-values are shown above each dataset.

### Selective deficit in FcγR3 binding to BA.2

Given the diminished binding profiles to the BA.2 variant, we next aimed to investigate whether this compromised recognition of the BA.2 spike also translated to reduced Fcγ-receptor (FcγR) binding. Thus, we profiled BNT162b2 induced Spike-specific FcγR binding across the 4 low-affinity human FcγRs involved in driving non-neutralizing Fc-effector functions (**Figure 2)**. Interestingly, binding to the opsonophagocytic activating FcγR2a showed diminished omicron BA.1 and BA.2 reactivity compared to the other to the D614G/wildtype, alpha, beta, and delta variants (**Figure 2A)**. Attenuated omicron BA.1 and BA.2 reactivity persisted with waning and after the boost, however FcγR2a binding to the BA.2 was not different at post vaccine timepoints (prime and boost) and remained higher compared to BA.1 at the waning timepoint. Conversely, the inhibitory FcγR2b was not different across the omicron sublineages at peak primary immunogenicity, BA.2 FcγR2b binding was higher than BA.1 at the waning timepoint, and BA.2 FcγR2b binding was lower than BA.1 binding after the boost, albeit omicron reactivity being lower than the other VOCs across all timepoints (**Figure 2B)**. Finally, vaccine induced Spike-binding antibodies to the cytotoxicity FcγR3a and neutrophil specific FcγR3b receptors showed a similar pattern of lower overall omicron binding responses after the peak primary immune response, although BA.2 exhibited the lowest FcγR3 binding (**Figure 2C-D)**. At waning timepoints both omicron responses were significantly lower than the other VOCs. After boosting, all VOC-FcγR3 binding responses increased, although omicron responses increased to a lesser degree, with BA.2 exhibiting the most compromised increase in FcγR3 binding. Thus, these data point to highly conserved activating opsonophagocytic inducing BA.2-antibody specific FcγR2a binding compared to BA.1, potentially continuing to attenuate disease, but a marked loss of FcγR2b, FcγR3a, and FcγR3b binding to the BA.2 sublineage resulting in compromised protection from transmission.

**Figure 2.**
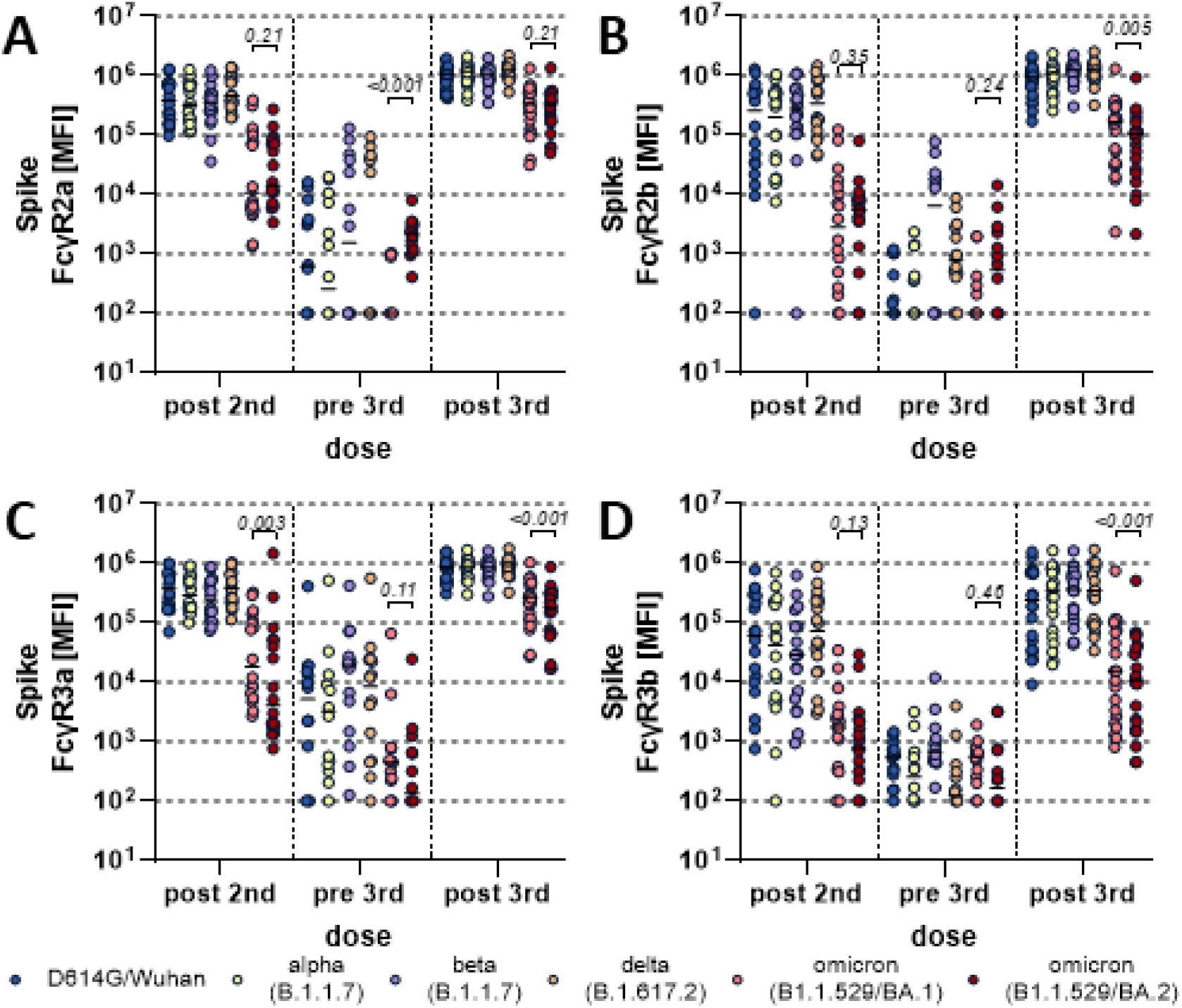
Fcγ-receptor binding antibody profiles across SARS-CoV-2 variants of concern. Individuals received a three-dose regimen of the BNT162b2 vaccine. Samples were taken approx. 4 weeks after the second dose (post 2nd, n=18), before the third dose approx. 8 months after the second dose (pre 3rd, n=14) and approx. 4 weeks after the third dose (post 3rd, n=22). Binding to FcγR2a (A), FcγR2b (B), FcγR3a (C) and FcγR3b (D) of D614G (WT), Alpha (B.1.1.7), Beta (B.1.351), Delta (B.1.617.2) variants of concern, and Omicron (B.1.1.529) BA.1 and BA.2 subvariants Spike specific antibodies were measured by Luminex. The average value of technical replicates is shown. The data was corrected for background and negative values were set to 100 for graphing purposes. A two-sided paired Wilcoxon test with a Benjamini-Hochberg post-test correcting for multiple comparisons was used to test for statistical differences between BA1 and BA2 titers, respectively. P-values are shown above each dataset.

### Vaccine induced antibodies poorly leverage neutrophil opsonophagocytosis against the BA.2 sublineage Spike

Given the observed compromised FcR binding profiles, we aimed to define whether vaccine induced antibodies could leverage antibody effector functions. At peak immunogenicity after the primary series, BNT162b2 induced Spike-specific antibodies drove significantly (p<0.001) compromised omicron BA.1 antibody-dependent neutrophil phagocytosis (ADNP) compared to the D614G/Wuhan Spike (**Figure 3**). Conversely, BNT162b2 induced Spike-specific functional activity was further compromised to the omicron BA.2 spike. ADNP activity declined over time proportionally to all of the Spike variants, albeit the BA.2 ADNP activity remained the lowest. Boosting increased ADNP activity to all Spike, although omicron Spike responses never reached those of the D614G/Wuhan response, and BA.2 remained the lowest. These data suggest that non-neutralizing antibody effector functions are severely compromised at all timepoints for the BA.2 omicron sublineage, likely resulting in compromised opsonization, FcR activation, and viral clearance to help attenuate disease and transmission.

**Figure 3.**
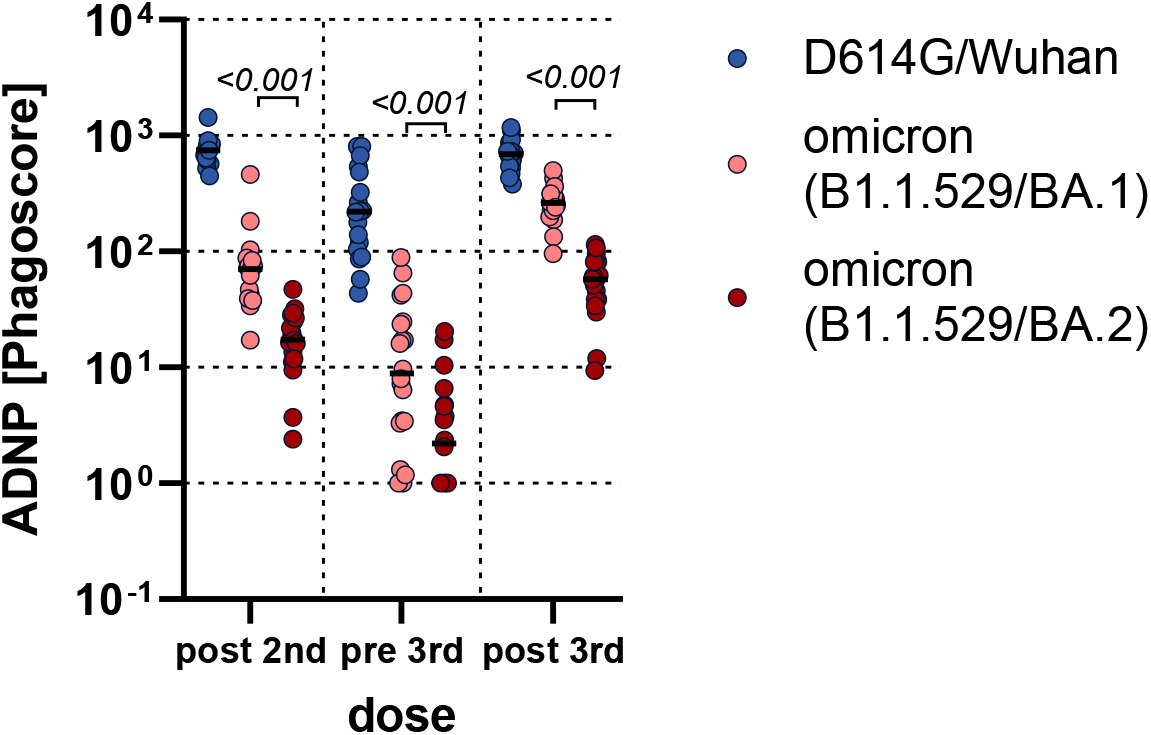
Fc functionality of BNT162b2 induced antibodies is superior to BA.1 compared to BA.2 antigen. Antibody-dependent Neutrophil Phagocytosis (ADNP) of D614G, Omicron (B1.1.529) BA.1 or BA.2 specific antibodies was analyzed in BNT162b2 recipients at approx. 4 weeks after the second dose (post 2nd, n=18), before the third dose approx. 8 month after the second dose (pre 3rd, n=14) and approx. 4 weeks after the third dose (post 3rd, n=22). The average value of two donors is shown. The data was corrected for background and negative values were set to 1 for graphing purposes. A two-sided paired Wilcoxon test with a Benjamini-Hochberg post-test correcting for multiple comparisons was used to test for statistical differences between BA1 and BA2 titers, respectively. P-values are shown above each dataset.

### Multivariate mucosal signatures of defective immunity to BA.2

To finally gain a detailed understanding of the fundamental differences in the BNT162b2 induced response to BA.1 and BA.2 that may result in less protection against BA.2, we generated a multivariate Partial least squares-discriminant analysis (PLS-DA) model across the 2 spike antigens at peak immunogenicity after the third dose (**Figure 4**). Perfect separation was noted across the BA.1 and BA.2 responses (**Figure 4A**), marked by elevated antibody-responses nearly exclusively to the BA.1 lineage. Specifically, BA.1 specific immune profiles exhibited a selective enrichment of Spike-specific Fcα-receptor binding antibodies, higher levels of Spike-specific IgA1 levels, enhanced ADNP, and elevated levels of Spike-IgG3 and FcγR2a levels (**Figure 4B**). Thus, beyond the overall compromised vaccine induced immune response to omicron BA.1, vaccine induced immune responses are further deficient to the BA.2, marked by more limited mucosal (IgA/FcαR) and IgG/FcγR responses that may lead to poorer control and clearance of this sublineage.

**Figure 4.**
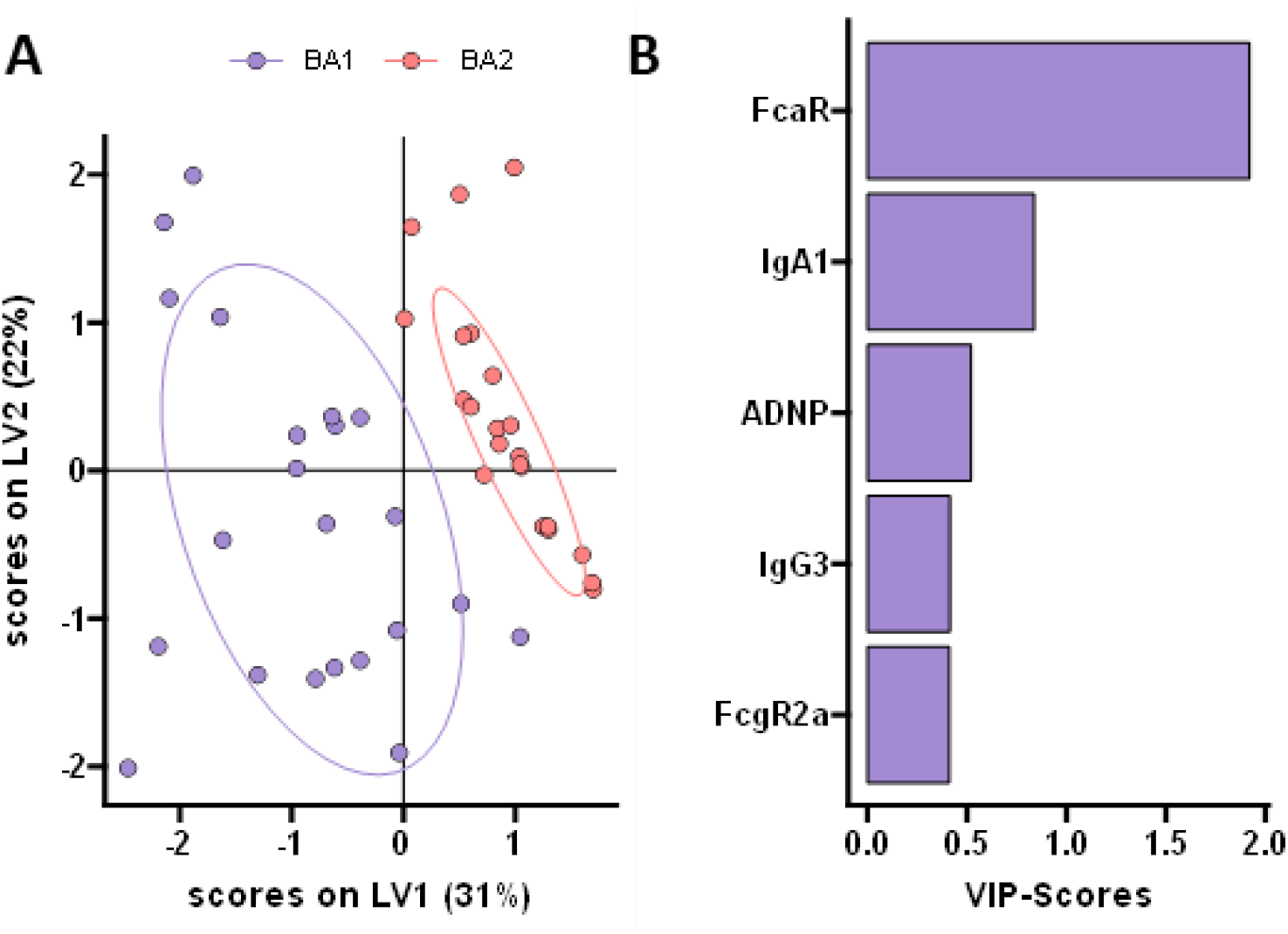
BNT162b2 induces distinct BA.1 and BA.2 specific responses. A machine learning model was built using BA.1 and BA.2 Spike specific features in BNT162b2 vaccinated individuals post third dose (n=22). A) A minimal set of LASSO selected features was used to discriminate between humoral responses in a PLS-DA analysis. Each point represents an individual’s humoral response for BA.1 (purple) and BA.2 (red). B) Selected features were ordered according to their Variable Importance in Projection (VIP) score (purple = enriched for BA.1 antigen).

## Discussion

Early Phase 3 vaccine trial immune correlates analyses, at a time when DG14G/Wuhan and the alpha variant dominated the global pandemic, pointed to the importance of neutralizing antibodies in protection against COVID-19(*17*). However, the emergence of the beta, delta and now omicron variants of concern, that significantly evade neutralizing antibody responses but do not cause more disease among vaccinated populations (*5-8*), has raised the possibility that alternative vaccine induced immune responses may be key to protection. Both vaccine induced T cell and non-neutralizing antibody responses have shown enhanced resilience to VOCs and have been proposed to contribute to attenuated disease against these evolving neutralizing antibody-escaping variants. However, the emergence of the omicron BA.2 sublineage has begun to challenge this hypothesis. Specifically, BA.2 exhibits enhanced transmissibility compared to the BA.1 lineage, despite only 1.5-fold reduced neutralizing antibody activity to BA.2 (*12*). Thus, reduced neutralizing antibody cannot account for enhanced transmissibility of this new sublineage of omicron. Given our increasing appreciation for a role for non-neutralizing Fc-effector antibody responses in protection against natural severe disease(*18, 19*), in monoclonal therapeutic activity (*13-16*), and in vaccine mediated protection(*20*), here we profiled the Pfizer BNT162b2 functional humoral immune response across VOCs including BA.1 and BA.2. As expected, vaccine induced non-neutralizing antibody responses were lower to omicron variants but were most profoundly diminished against the BA.2 Spike antigen. However, selective Fc-receptor binding deficits were noted, marked by maintained/superior FcγR2a/b binding, but reduced FcγR3a/3b binding to BA.2 compared to BA.1. Linked to reduced neutrophil opsonophagocytic activity, these data point to a selective loss of IgA/neutrophil functional mucosal immunity, that may be key to protection against transmission, but maintenance of FcγR2a/b that may continue to confer protection against disease.

Omicron BA.1 included 36 mutations in the Spike antigen that significantly evaded vaccine induced and monoclonal therapeutic recognition (*5*). Similarly, to the loss of neutralizing antibody responses, Moderna mRNA1273, Pfizer BNT162b2, and CoronaVac vaccine induced immune responses lost recognition of RBD but maintained robust recognition and non-neutralizing antibody responses to the BA.1 spike antigen (*21*). Conversely, BA.2 includes 11 additional Spike mutations that appear to further compromise antibody binding to the Spike, resulting in compromised Fc-receptor binding, and result in significantly lower non-neutralizing antibody responses to this new sublineage. Whether these additional 11 mutations result in disruption of immune complex density, or create geometric changes in the spike, precluding the formation of robust immune complexes and binding to particular Fc-receptors remains incompletely understood but could provide key insights for the design of next generation vaccine-antigen inserts to maximize the induction of highly functional antibodies that, in addition to neutralization, may be key to protection against transmission.

Direct comparison of BNT162b2 immunity to BA.1 and BA.2 pointed to a selective deficit of IgA, IgG1, Fc-receptor, and neutrophil phagocytic responses targeting the BA.2 spike. These four vaccine induced antibody features point to a unique axis of immunity that may be lost to BA.2. Specifically, neutrophils respond to both IgG1/3 immune complexes via antibody-binding to FcγR2a and FcγR3b as well as to IgA-formed immune complexes via the constitutive expression of FcαR (*22, 23*), enabling neutrophils to rapidly contain immune complexes both systemically as well as at mucosal membranes. Thus, the selective loss of IgA/FcαR/ADNP activity against BA.2 may point to a selective deficit in mucosal immunity, resulting in increased transmission as well as enhanced respiratory disease related to the selective evasion of this variant.

Emerging vaccine efficacy data point to preserved protection against BA.2 (*24*). However, why similar levels of neutralization permit BA.2 to infect more efficiently than BA.1 may both be related to changes in the viral spike antigen(*25*), but also related to compromised alternative antibody effector functions that may work synergistically with neutralization to promote full control of the virus at the mucosal barrier. Opsonophagocytic functions at the mucosal barrier are linked to protection against several infectious diseases including Streptococcus pneumoniae (*26*), RSV (*27*), Influenza (*28*), etc.. Moreover, Fc-effector functions may potentiate the immune-protective role of neutralizing antibodies, via a collaboration between the constant-domain (Fc) and antigen binding (Fab) domain of antibodies. Thus, it is plausible that the selective loss of collaborative IgA/IgG opsonophagocytosis may diminish the initial barrier of protection against BA.2, despite relatively preserved neutralization, pointing to opsonophagocytosis, coupled to neutralization, as potential critical correlate of immunity against transmission.

While this study did not explore functional humoral immunity to BA.2 following additional vaccine platforms, increased BA.2 transmission appears to occur globally in a vaccine platform independent manner. The additional evolution of 11 mutations in the BA.2 sublineage of omicron does not appreciably evade neutralization, but does specifically subvert mucosal non-neutralizing humoral immunity, potentially resulting in significantly enhanced transmission. Several immunological mechanisms have been proposed as key correlates of immunity against COVID-19, including a role for neutralizing antibodies, T cells, memory B cells, and non-neutralizing antibody responses. However, each of these arms of the immune response do not function alone, and instead function as a collective in the response to evolving pathogens, working to complement one another to achieve rapid control of infection. Thus, while neutralizing antibodies and T cell immune responses have traditionally been used as correlates of immunity to advance vaccine development, emerging data from real-world vaccine rollout, in the face of evolving SARS-CoV-2 variants, points to a critical role for non-neutralizing antibody responses in attenuation of disease. Thus, contemporized vaccine design and evaluation, able to capture changes to Fc-effector functions, may help with the design of next generation vaccines able to provide protection against current and future SARS-CoV-2 variants.

## Data Availability

All data produced in the present study are available upon reasonable request to the authors.

## Acknowledgments

We thank Nancy Zimmerman, Mark and Lisa Schwartz, an anonymous donor (financial support), Terry and Susan Ragon, and the SAMANA Kay MGH Research Scholars award for their support. We acknowledge support from the Ragon Institute of MGH, MIT, and Harvard, the Massachusetts Consortium on Pathogen Readiness (MassCPR) and the Musk foundation. All data associated with this study are in the paper or supplementary materials.

## Author contributions

Y.C.B. and J.K. performed the experiments. Y.C.B., D.C. and G.A. analyzed and interpreted the data. H.G., W.S., and B.C. expressed and purified protein antigens. A.C., A.Y.C., and D.H.B supervised and managed the sample collection. G.A. supervised the project. Y.C.B. and G.A. drafted the manuscript. All authors critically reviewed the manuscript.

## Methods

### Study population

Plasma samples from BNT162b2 vaccinated individuals (median age: 34 years, range: 23-69 years, 88 % female) were obtained from a specimen biorepository at Beth Israel Deaconess Medical Center (BIDMC). Participants received three doses of 30 μg BNT162b2. The first doses were given approx. 21 days apart as per manufacturers recommendation. The third dose was given a median 254 days (range 248-258 days) after the second dose. All participants provided informed consent prior to enrollment into the study. No participant reported previous SARS-CoV-2 infection, received other COVID-19 vaccines, or immunosuppressive medications. This study was overseen and approved by the BIDMC Institutional Review Board (#2020P000361) and the MassGeneral Institutional Review Board (#2021P002628).

### Antigens

Spike protein antigens for the D614G wildtype, alpha (B.1.1.7), beta (B.1.351), and delta (B.1.617.2) VOCs were obtained from Sino-Biologicals. Omicron (B.1.1.529) BA1 and BA2 Spikes were produced in house. All antigens were produced in mammalian HEK293 cells.

### IgG subclass, isotype and FcγR binding

Antigen specific antibody subclass, isotypes, and FcγR binding was analyzed in technical replicates by Luminex technology. Antigens were coupled to Luminex beads (Luminex Corp, TX, USA) by carbodiimide-NHS ester-chemistry with an individual region per antigen. Coupled beads were incubated with diluted plasma sample (1:100 for IgG3, IgM and IgA1, 1:500 for IgG1, FcαR and 1:2,000 for FcγR probing) for 2 hours at room temperature in 384 well plates (Greiner Bio-One, Germany). Unbound antibodies were washed away and subclasses, isotypes were detected with a respective PE-conjugated antibody (anti-human IgG1 (Cat# 9052-09, RRID:AB_2796621), IgG3 (Cat# 9210-09, RRID:AB_2796701), IgM (Cat# 9020-09, RRID:AB_2796577) or IgA1 (Cat# 9130-09, RRID:AB_2796656) all SouthernBiotech, AL, USA) at a 1:100 dilution. For the analysis of FcγR binding PE-Streptavidin (Agilent Technologies, CA, USA) was coupled to recombinant and biotinylated human FcγR2a, FcγR2b, FcγR3a, FcγR3b or FcαR protein. Coupled FcR were used as a secondary probe at a 1:1000 dilution. After 1 h incubation, excessive secondary reagent was washed away and the relative antibody concentration per antigen determined on an IQue analyzer (IntelliCyt).

### Antibody-Dependent-Neutrophil-Phagocytosis (ADNP)

Phagocytosis score of primary human neutrophils was determined as described before (*23*). Antigens were biotinylated with NHS-Sulfo-LC-LC kit according to the manufacturer’s instruction (Thermo Fisher). Excessive biotin was removed by size exclusion chromatography using Zeba-Spin desalting columns (7kDa cutoff, Thermo Fisher). Biotinylated antigens were coupled to fluorescent neutravidin beads (Thermo Fisher) and incubated with 1:10 diluted plasma. Primary cells were derived from Ammonium-Chloride-Potassium (ACK) buffer lysed whole blood from healthy donors and incubated with immune complexes for 1h at 37°C. Neutrophils were stained for surface CD66b (Biolegend, CA, USA; clone: G10F5) expression, fixed with 4% para-formaldehyde and analyzed an IQue analyzer (IntelliCyt).

### Computational analysis

A multivariate classification model was built to discriminate humoral profiles between BA.1 and BA.2 specific antibodies. Prior to analysis, all data were normalized using z-scoring. Feature selection was performed using least absolute shrinkage and selection operator (LASSO). Classification and visualization were performed using partial least square discriminant analysis (PLS-DA). Selected features were ordered according to their Variable Importance in Projection (VIP) score and the first two latent variables (LVs) of the PLS-DA model were used to visualize the samples. These analyses were performed using R package “ropls” version 1.20.0 (*29*) and “glmnet” version 4.0.2 (*30*) and the systemseRology R package (v.1.1) (https://github.com/LoosC/systemsseRology).

### Statistical analysis

If not stated otherwise, we assumed non-normal distributions and plots were generated and statistical differences between two groups were calculated in Graph Pad Prism V.9. A paired Wilcoxon test with a Benjamini-Hochberg post-test correcting for multiple comparisons was used to test for statistical differences between BA1 and BA2 features at the different timepoints.

## References

1. L. Ulrich, N. J. Halwe, A. Taddeo, N. Ebert, J. Schön, C. Devisme, B. S. Trüeb, B. Hoffmann, M. Wider, X. Fan, M. Bekliz, M. Essaidi-Laziosi, M. L. Schmidt, D. Niemeyer, V. M. Corman, A. Kraft, A. Godel, L. Laloli, J. N. Kelly, B. M. Calderon, A. Breithaupt, C. Wylezich, I. Berenguer Veiga, M. Gultom, S. Osman, B. Zhou, K. Adea, B. Meyer, C. S. Eberhardt, L. Thomann, M. Gsell, F. Labroussaa, J. Jores, A. Summerfield, C. Drosten, I. A. Eckerle, D. E. Wentworth, R. Dijkman, D. Hoffmann, V. Thiel, M. Beer, C. Benarafa, Enhanced fitness of SARS-CoV-2 variant of concern Alpha but not Beta. Nature 602, 307–313 (2022); published online Epub2022/02/01 (10.1038/s41586-021-04342-0).

2. J. Zahradník, S. Marciano, M. Shemesh, E. Zoler, D. Harari, J. Chiaravalli, B. Meyer, Y. Rudich, C. Li, I. Marton, O. Dym, N. Elad, M. G. Lewis, H. Andersen, M. Gagne, R. A. Seder, D. C. Douek, G. Schreiber, SARS-CoV-2 variant prediction and antiviral drug design are enabled by RBD in vitro evolution. Nature Microbiology 6, 1188–1198 (2021); published online Epub2021/09/01 (10.1038/s41564-021-00954-4).

3. B. J. Willett, J. Grove, O. A. MacLean, C. Wilkie, N. Logan, G. D. Lorenzo, W. Furnon, S. Scott, M. Manali, A. Szemiel, S. Ashraf, E. Vink, W. T. Harvey, C. Davis, R. Orton, J. Hughes, P. Holland, V. Silva, D. Pascall, K. Puxty, A. da Silva Filipe, G. Yebra, S. Shaaban, M. T. G. Holden, R. M. Pinto, R. Gunson, K. Templeton, P. R. Murcia, A. H. Patel, T. C.-G. U. Consortium, J. Haughney, D. L. Robertson, M. Palmarini, S. Ray, E. C. Thomson, The hyper-transmissible SARS-CoV-2 Omicron variant exhibits significant antigenic change, vaccine escape and a switch in cell entry mechanism. medRxiv, 2022.2001.2003.21268111 (2022)10.1101/2022.01.03.21268111).

4. W. T. Harvey, A. M. Carabelli, B. Jackson, R. K. Gupta, E. C. Thomson, E. M. Harrison, C. Ludden, R. Reeve, A. Rambaut, S. J. Peacock, D. L. Robertson, C.-G. U. Consortium, SARS-CoV-2 variants, spike mutations and immune escape. Nature Reviews Microbiology 19, 409–424 (2021); published online Epub2021/07/01 (10.1038/s41579-021-00573-0).

5. L. A. VanBlargan, J. M. Errico, P. J. Halfmann, S. J. Zost, J. E. Crowe, L. A. Purcell, Y. Kawaoka, D. Corti, D. H. Fremont, M. S. Diamond, An infectious SARS-CoV-2 B.1.1.529 Omicron virus escapes neutralization by several therapeutic monoclonal antibodies. bioRxiv, 2021.2012.2015.472828 (2021)10.1101/2021.12.15.472828).

6. T. P. Peacock, J. C. Brown, J. Zhou, N. Thakur, J. Newman, R. Kugathasan, K. Sukhova, M. Kaforou, D. Bailey, W. S. Barclay, The SARS-CoV-2 variant, Omicron, shows rapid replication in human primary nasal epithelial cultures and efficiently uses the endosomal route of entry. bioRxiv, 2021.2012.2031.474653 (2022)10.1101/2021.12.31.474653).

7. K. McMahan, V. Giffin, L. H. Tostanoski, B. Chung, M. Siamatu, M. S. Suthar, P. Halfmann, Y. Kawaoka, C. Piedra-Mora, A. J. Martinot, S. Kar, H. Andersen, M. G. Lewis, D. H. Barouch, Reduced Pathogenicity of the SARS-CoV-2 Omicron Variant in Hamsters. bioRxiv, 2022.2001.2002.474743 (2022)10.1101/2022.01.02.474743).

8. S. Collie, J. Champion, H. Moultrie, L. G. Bekker, G. Gray, Effectiveness of BNT162b2 Vaccine against Omicron Variant in South Africa. N Engl J Med, (2021); published online EpubDec 29 (10.1056/NEJMc2119270).

9. D. Yamasoba, I. Kimura, H. Nasser, Y. Morioka, N. Nao, J. Ito, K. Uriu, M. Tsuda, J. Zahradnik, K. Shirakawa, R. Suzuki, M. Kishimoto, Y. Kosugi, K. Kobiyama, T. Hara, M. Toyoda, Y. L. Tanaka, E. P. Butlertanaka, R. Shimizu, H. Ito, L. Wang, Y. Oda, Y. Orba, M. Sasaki, K. Nagata, K. Yoshimatsu, H. Asakura, M. Nagashima, K. Sadamasu, K. Yoshimura, J. Kuramochi, M. Seki, R. Fujiki, A. Kaneda, T. Shimada, T.-a. Nakada, S. Sakao, T. Suzuki, T. Ueno, A. Takaori-Kondo, K. J. Ishii, G. Schreiber, T. G. t. P. J. Consortium, H. Sawa, A. Saito, T. Irie, S. Tanaka, K. Matsuno, T. Fukuhara, T. Ikeda, K. Sato, Virological characteristics of SARS-CoV-2 BA.2 variant. bioRxiv, 2022.2002.2014.480335 (2022)10.1101/2022.02.14.480335).

10. F. P. Lyngse, C. T. Kirkeby, M. Denwood, L. E. Christiansen, K. Mølbak, C. H. Møller, R. L. Skov, T. G. Krause, M. Rasmussen, R. N. Sieber, T. B. Johannesen, T. Lillebaek, J. Fonager, A. Fomsgaard, F. T. Møller, M. Stegger, M. Overvad, K. Spiess, L. H. Mortensen, Transmission of SARS-CoV-2 Omicron VOC subvariants BA.1 and BA.2: Evidence from Danish Households. medRxiv, 2022.2001.2028.22270044 (2022)10.1101/2022.01.28.22270044).

11. S. F. Ahmed, A. A. Quadeer, M. R. McKay, SARS-CoV-2 T cell responses are expected to remain robust against Omicron. bioRxiv, 2021.2012.2012.472315 (2021)10.1101/2021.12.12.472315).

12. J. Yu, A.-r. Y. Collier, M. Rowe, F. Mardas, J. D. Ventura, H. Wan, J. Miller, O. Powers, B. Chung, M. Siamatu, N. P. Hachmann, N. Surve, F. Nampanya, A. Chandrashekar, D. H. Barouch, Comparable Neutralization of the SARS-CoV-2 Omicron BA.1 and BA.2 Variants. medRxiv, 2022.2002.2006.22270533 (2022)10.1101/2022.02.06.22270533).

13. C. E. Z. Chan, S. G. K. Seah, H. Chye, S. Massey, M. Torres, A. P. C. Lim, S. K. K. Wong, J. J. Y. Neo, P. S. Wong, J. H. Lim, G. S. L. Loh, D. Wang, J. D. Boyd-Kirkup, S. Guan, D. Thakkar, G. H. Teo, K. Purushotorman, P. E. Hutchinson, B. E. Young, J. G. Low, P. A. MacAry, H. Hentze, V. S. Prativadibhayankara, K. Ethirajulu, J. E. Comer, C. K. Tseng, A. D.T. Barrett, P. J. Ingram, T. Brasel, B. J. Hanson, The Fc-mediated effector functions of a potent SARS-CoV-2 neutralizing antibody, SC31, isolated from an early convalescent COVID-19 patient, are essential for the optimal therapeutic efficacy of the antibody. PLoS One 16, e0253487 (2021)10.1371/journal.pone.0253487).

14. A. Schafer, F. Muecksch, J. C. C. Lorenzi, S. R. Leist, M. Cipolla, S. Bournazos, F. Schmidt, R. M. Maison, A. Gazumyan, D. R. Martinez, R. S. Baric, D. F. Robbiani, T. Hatziioannou, J. V. Ravetch, P. D. Bieniasz, R. A. Bowen, M. C. Nussenzweig, T. P. Sheahan, Antibody potency, effector function, and combinations in protection and therapy for SARS-CoV-2 infection in vivo. J Exp Med 218, (2021); published online EpubMar 1 (10.1084/jem.20201993).

15. E. S. Winkler, P. Gilchuk, J. Yu, A. L. Bailey, R. E. Chen, Z. Chong, S. J. Zost, H. Jang, Y. Huang, J. D. Allen, J. B. Case, R. E. Sutton, R. H. Carnahan, T. L. Darling, A. C. M. Boon, M. Mack, R. D. Head, T. M. Ross, J. E. Crowe, Jr., M. S. Diamond, Human neutralizing antibodies against SARS-CoV-2 require intact Fc effector functions for optimaltherapeutic protection. Cell 184, 1804–1820 e1816 (2021); published online EpubApr 1 (10.1016/j.cell.2021.02.026).

16. I. Ullah, J. Prevost, M. S. Ladinsky, H. Stone, M. Lu, S. P. Anand, G. Beaudoin-Bussieres, K. Symmes, M. Benlarbi, S. Ding, R. Gasser, C. Fink, Y. Chen, A. Tauzin, G. Goyette, C. Bourassa, H. Medjahed, M. Mack, K. Chung, C. B. Wilen, G. A. Dekaban, J. D. Dikeakos, E. A. Bruce, D. E. Kaufmann, L. Stamatatos, A. T. McGuire, J. Richard, M. Pazgier, P. J. Bjorkman, W. Mothes, A. Finzi, P. Kumar, P. D. Uchil, Live imaging of SARS-CoV-2 infection in mice reveals that neutralizing antibodies require Fc function for optimal efficacy. Immunity 54, 2143–2158 e2115 (2021); published online EpubSep 14 (10.1016/j.immuni.2021.08.015).

17. D. S. Khoury, D. Cromer, A. Reynaldi, T. E. Schlub, A. K. Wheatley, J. A. Juno, K. Subbarao, S. J. Kent, J. A. Triccas, M. P. Davenport, Neutralizing antibody levels are highly predictive of immune protection from symptomatic SARS-CoV-2 infection. Nat Med 27, 1205–1211 (2021); published online EpubJul (10.1038/s41591-021-01377-8).

18. T. Zohar, C. Loos, S. Fischinger, C. Atyeo, C. Wang, M. D. Slein, J. Burke, J. Yu, J. Feldman, B. M. Hauser, T. Caradonna, A. G. Schmidt, Y. Cai, H. Streeck, E. T. Ryan, D. H. Barouch, R. C. Charles, D. A. Lauffenburger, G. Alter, Compromised Humoral Functional Evolution Tracks with SARS-CoV-2 Mortality. Cell 183, 1508–1519 e1512 (2020); published online EpubDec 10 (10.1016/j.cell.2020.10.052).

19. O. S. Adeniji, L. B. Giron, M. Purwar, N. F. Zilberstein, A. J. Kulkarni, M. W. Shaikh, R. A. Balk, J. N. Moy, C. B. Forsyth, Q. Liu, H. Dweep, A. Kossenkov, D. B. Weiner, A. Keshavarzian, A. Landay, M. Abdel-Mohsen, COVID-19 Severity Is Associated with Differential Antibody Fc-Mediated Innate Immune Functions. mBio 12, (2021); published online EpubApr 20 (10.1128/mBio.00281-21).

20. M. J. Gorman, N. Patel, M. Guebre-Xabier, A. L. Zhu, C. Atyeo, K. M. Pullen, C. Loos, Y. Goez-Gazi, R. Carrion, Jr., J. H. Tian, D. Yuan, K. A. Bowman, B. Zhou, S. Maciejewski, M. E. McGrath, J. Logue, M. B. Frieman, D. Montefiori, C. Mann, S. Schendel, F. Amanat, F. Krammer, E. O. Saphire, D. Lauffenburger, A. M. Greene, A. D. Portnoff, M. J. Massare, L. Ellingsworth, G. Glenn, G. Smith, G. Alter, Fab and Fc contribute to maximal protection against SARS-CoV-2 following NVX-CoV2373 subunit vaccine with Matrix-M vaccination. Cell Rep Med, 100405 (2021); published online EpubAug 31 (10.1016/j.xcrm.2021.100405).

21. Y. Bartsch, X. Tong, J. Kang, M. J. Avendaño, E. F. Serrano, T. García-Salum, C. Pardo-Roa, A. Riquelme, R. A. Medina, G. Alter, Preserved Omicron Spike specific antibody binding and Fc-recognition across COVID-19 vaccine platforms. medRxiv, 2021.2012.2024.21268378 (2021)10.1101/2021.12.24.21268378).

22. J. E. Bakema, M. van Egmond, The human immunoglobulin A Fc receptor FcalphaRI: a multifaceted regulator of mucosal immunity. Mucosal Immunol 4, 612–624 (2011); published online EpubNov (10.1038/mi.2011.36).

23. C. B. Karsten, N. Mehta, S. A. Shin, T. J. Diefenbach, M. D. Slein, W. Karpinski, E. B. Irvine, T. Broge, T. J. Suscovich, G. Alter, A versatile high-throughput assay to characterize antibody-mediated neutrophil phagocytosis. J Immunol Methods 471, 46–56 (2019); published online EpubAug (10.1016/j.jim.2019.05.006).

24. U. H. S. Agency, COVID-19 vaccine surveillance report: 24 February 2022 (week 8). (2022); published online Epub02/24/2022 (

25. S. Kumar, K. Karuppanan, G. Subramaniam, Omicron (BA.1) and Sub-Variants (BA.1, BA.2 and BA.3) of SARS-CoV-2 Spike Infectivity and Pathogenicity: A Comparative Sequence and Structural-based Computational Assessment. bioRxiv, 2022.2002.2011.480029 (2022)10.1101/2022.02.11.480029).

26. S. Romero-Steiner, C. E. Frasch, G. Carlone, R. A. Fleck, D. Goldblatt, M. H. Nahm, Use of opsonophagocytosis for serological evaluation of pneumococcal vaccines. Clin Vaccine Immunol 13, 165–169 (2006); published online EpubFeb (10.1128/CVI.13.2.165-169.2006).

27. T. Zohar, J. C. Hsiao, N. Mehta, J. Das, A. Devadhasan, W. Karpinski, C. Callahan, M. P. Citron, D. J. DiStefano, S. Touch, Z. Wen, J. R. Sachs, P. J. Cejas, A. S. Espeseth, D. A. Lauffenburger, A. J. Bett, G. Alter, Upper and lower respiratory tract correlates of protection against respiratory syncytial virus following vaccination of nonhuman primates. Cell Host Microbe 30, 41–52 e45 (2022); published online EpubJan 12 (10.1016/j.chom.2021.11.006).

28. V. C. Huber, J. M. Lynch, D. J. Bucher, J. Le, D. W. Metzger, Fc receptor-mediated phagocytosis makes a significant contribution to clearance of influenza virus infections. J Immunol 166, 7381–7388 (2001); published online EpubJun 15 (10.4049/jimmunol.166.12.7381).

29. E. A. Thevenot, A. Roux, Y. Xu, E. Ezan, C. Junot, Analysis of the Human Adult Urinary Metabolome Variations with Age, Body Mass Index, and Gender by Implementing a Comprehensive Workflow for Univariate and OPLS Statistical Analyses. J Proteome Res 14, 3322–3335 (2015); published online EpubAug 7 (10.1021/acs.jproteome.5b00354).

30. J. H. Friedman, T. Hastie, R. Tibshirani, Regularization Paths for Generalized Linear Models via Coordinate Descent. 2010 33, 22 (2010); published online Epub2010-02-02 (10.18637/jss.v033.i01).

